# Analysis of Extraocular muscle volumes in idiopathic Hypertrophic Pachymeningitis patients

**DOI:** 10.1101/2024.08.18.24312196

**Authors:** Suppakul Kitkamolwat, Supichaya Soonthornpusit, Akarawit Eiamsamarng, Natthapon Rattanathamsakul, Niphon Chirapapaisan, Chanon Ngamsombat

**Affiliations:** Department of Radiology, Faculty of Medicine Siriraj Hospital, Mahidol University, Bangkok, Thailand; Department of Ophthalmology, Faculty of Medicine Siriraj Hospital, Mahidol University, Bangkok, Thailand; Department of Medicine, Faculty of Medicine Siriraj Hospital, Mahidol University, Bangkok, Thailand

**Keywords:** idiopathic pachymeningitis, extraocular muscle, volume measurement

## Abstract

**Background:** Idiopathic hypertrophic pachymeningitis (HP) is a rare chronic inflammatory condition without an identifiable cause characterized by fibrous thickening of the dura mater, which can involve the extraocular muscles (EOM).

**Objective:** To evaluate volumetric changes of EOM in idiopathic HP patients compared with healthy controls (HC) and study the correlation with ocular motility disturbance.

**Materials and methods:** A retrospective study of 22 diagnosed idiopathic HP patients and 22 age-matched, sex-matched HC underwent a 3T MRI scan from January 1, 2017, to December 31, 2022. EOM was manually segmented from the T1W image using 3D Slicer software, and volume was calculated using FSL software. T-tests and Mann-Whitney U tests were used to compare EOM volumes between the idiopathic HP and control groups. Pearson’s correlation coefficient was then used to assess the correlation between ocular motility and EOM enlargement.

**Results:** In idiopathic HP patients, the average EOM volumes, including the medial rectus (MR), inferior rectus (IR), inferior oblique (IO), right lateral rectus (LR), right superior oblique (SO), and left superior rectus (SR) muscles, were significantly larger compared to those in HC, particularly in the left IR and both MR. However, there was no significant correlation between the enlargement of these 9 EOMs and the extraocular movement limitation.

**Conclusion:** In idiopathic HP patients, significantly larger EOM volumes were found compared to control subjects. This enlargement could be due to the diffuse infiltrative histopathology potentially involving microstructures in the EOM. Extraocular movement limitations may be related to cranial nerve involvement.

## 1. Introduction

Hypertrophic pachymeningitis (HP) is a rare chronic inflammatory condition characterized by fibrous thickening of the cerebral and/or spinal dura mater. HP is classified into two types based on its etiology: primary (idiopathic) HP, which has no identifiable cause, and secondary HP, which includes coexisting causes such as infectious (such as tuberculosis, syphilis, and sarcoidosis), autoimmune (such as IgG4-related disease), vasculitis (such as Wegener’s granulomatosis), and neoplastic (such as lymphoma and metastasis).

Idiopathic HP is the most predominant etiology of HP (1-3). However, the pathophysiology of idiopathic HP is currently unclear. Histopathological examination reveals non-specific chronic inflammatory changes in the dura, including extensive fibrosis and often-detected inflammatory cell infiltration. The dura mater surface may contain small, mature lymphocytes, plasma cells, and epithelioid histiocytes (4).

HP is diagnosed based on clinical symptoms such as headaches, diplopia, visual loss, ptosis, and facial pain, with thickening and enhancement of the dura mater seen on MRI imaging and/or dura mater biopsy, which is the gold standard of diagnosis.

HP is a rare condition, with a reported prevalence of 0.949 per 100,000 people (5). A prior study examined the clinical presentation, laboratory findings, and neuroradiologic characteristics of HP. The results showed that idiopathic HP is the most common type, with diplopia being the most common clinical manifestation (85.2%). CN VI was found to be the most commonly involved cranial nerve, and the cavernous sinus was the most common location of cranial dural thickening (3).

The aim of our study was to evaluate the volumetric changes of all EOM in idiopathic HP patients compared to age-matched, sex-matched healthy controls (HC) and to study the correlation with ocular motility disturbance. Our hypothesis was that the abnormal EOM volumes cause extraocular muscle limitation.

## 2. Materials and methods

### Participants

This retrospective study included twenty-two patients diagnosed with idiopathic HP at Siriraj Hospital using search tools in the picture archiving and communication system (PACS). The Siriraj institutional review board approved the study (certificate of approval number Si 055/2023). The data was collected between January 1, 2017, and December 31, 2022.

The entry criteria were defined as follows: 1) The diagnosis was based on dura mater biopsy or dura mater thickening or enhancement on MRI imaging, and 2) dural thickening or enhancement could not be explained by intracranial hypotension, neoplastic pachymeningitis, or other coexisting conditions. (3)

Patients with a history of other etiologies affecting EOMs, such as prior brain surgery or trauma, underlying brain tumor or brain metastasis, focal mass at the EOM, or any EOM enlargement recorded in the radiologist’s official report, were excluded.

We included twenty-two age-matched and sex-matched healthy controls who underwent MRI for non-specific indications without eye symptoms and no underlying eye disease with normal MRI findings for comparison.

### Ophthalmological examination

The data was gathered from medical records recorded by ophthalmologists at Siriraj Hospital. EOM function was evaluated by asking the patient to move their eyes in an H-shaped pattern. The ophthalmic examination was performed by trained technicians.

### MRI image acquisition

All patients were scanned using clinical MRI scanners at Siriraj Hospital: Signa 3.0T (General Electric Healthcare, USA), MAGNETOM Vida 3.0T (Siemens Healthcare, Germany), and Archieva 3.0T (Philips Healthcare, Netherlands) with a 32-channel head coil. The MRI protocol included a T1W image sequence using three-dimensional TFE with the following parameters: echo time (TE) = 4.61 ms, repetition time (TR) = 9.53 ms, matrix size = 352 × 352, field-of-view (FOV) = 230 × 230 × 172 mm^3^, voxel size = 0.72 × 0.72 × 0.5 mm^3^, flip angle = 8°, scan time 6 minutes.

### Data processing

In brief, all EOM raw data were collected as DICOM images and transformed to NiFTI format using DICOM to NiFTI software.

Segmentation of the EOM was performed using 3D Slicer software v.5.6.1. The ROI of the EOM was drawn and finalized by a 3rd-year resident and a 15-year-experienced neuroradiologist. The medial rectus (MR), inferior rectus (IR), lateral rectus (LR), superior oblique (SO), and inferior oblique (IO) muscles were drawn separately in the coronal plane of the T1W image (Fig. 1). Considering their anatomical closeness, the superior rectus (SR) and levator palpebrae superioris muscles were measured together and named as the superior rectus muscle (6-9).

**Fig. 1.**
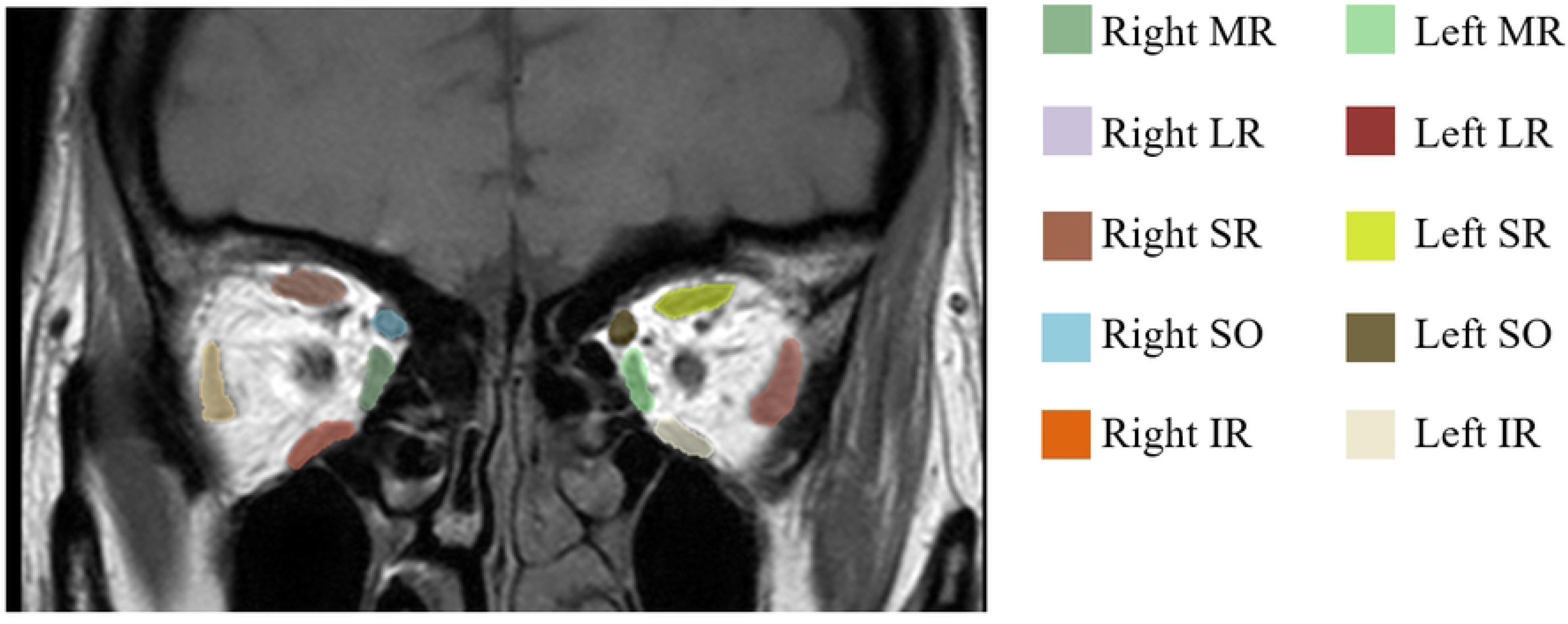
MRI T1 weighted images in coronal plane of a representative case with EOM segmentation by using 3D slicer software.

The calculation of each EOM volume from the outlined ROI was performed using FSL Software (https://fsl.fmrib.ox.ac.uk).

## 3. Statistic Analysis

After the image processing was done, statistical analysis was performed using SPSS (PASW Statistics for Windows, version 18.0).

Demographic data, including age, sex, and duration between eye and MRI examinations, were reported as means and standard deviations (SD). The quantitative data of the subjects with idiopathic HP and the healthy control group were compared using T-tests. The qualitative data of the subjects with idiopathic HP and the healthy control group were compared using the Chi-square test. Categorical data, including the location of dural thickening, were reported as frequency and percentage.

Comparisons of EOM volumes between patients and control groups were performed using T-tests for normal distributions and the Mann-Whitney U test for non-normal distributions. Pearson’s correlation was used to identify a significant correlation between EOM volume and ocular motility. The criteria for statistical significance were defined as a p-value < 0.05.

## 4. Results

### Clinical description

Clinical and demographic data for the idiopathic HP patients and HC are presented in Table 1. Twenty-two idiopathic HP patients, including 8 males and 14 females (mean age 51.50 ± 15.41 years), and 22 healthy controls in the sex-matched and age-matched groups (mean age 51.41 ± 15.30 years) were included for analysis. There is no statistical difference in age or sex between the idiopathic HP and control groups (p=0.984 and p=1.000, respectively). The mean duration between the date of eye examination and MRI examination is 59.64 ± 99.40 days.

**Table 1.** Demographic data.

|  | Group, n (%) |  | P-value |
| --- | --- | --- | --- |
|  | Idiopathic HP (n=22) | Healthy control (n=22) |  |
| Age (mean±sd) (years) | 51.50±15.41 | 51.41±15.30 | 0.984 |
| Sex |  |  | 1.000 |
| Male | 8 (36.4) | 8 (36.4) |  |
| Female | 14 (63.6) | 14 (63.6) |  |
| Duration between eye examination and MRI imaging (mean±sd) (days) | 59.64±99.40 | - |  |

In idiopathic HP patients, most of the dural thickening is involved in the cavernous sinus and tentorium cerebelli (each 11/22 patients; 50%).

### MRI imaging analysis

The total 12 EOM volumes from both eyes of idiopathic HP patients and healthy control groups were measured and summarized in Table 2.

**Table 2.** Statistic data compared the average extraocular muscle volume (mm^3^) of idiopathic HP patients versus healthy control patients.

| Extraocular muscle | EOM volume (mm<sup>3</sup>) |  | P-value |
| --- | --- | --- | --- |
|  | Idiopathic HP ;mean±SD | Healthy control ;mean±SD |  |
| Right Medial rectus (MR) | 560.65±122.89 | 445.47±111.97 | 0.002* |
| Right Lateral rectus (LR) | 624.03±131.76 | 532.82±99.77 | 0.005* |
| Right Superior rectus (SR) | 509.63±154.99 | 438.97±118.05 | 0.096 |
| Right Superior oblique (SO) | 340.71±70.15 | 271.07±81.23 | 0.004* |
| Right Inferior rectus (IR) | 496.78±134.01 | 403.97±82.60 | 0.008* |
| Right Inferior oblique (IO) | 300.31±117.56 | 217.45±79.83 | 0.009* |
| Left Medial rectus (MR) | 554.06±138.19 | 436.26±100.66 | 0.002* |
| Left Lateral rectus (LR) | 599.05±145.34 | 505.53±130.41 | 0.054 |
| Left Superior rectus (SR) | 533.22±146.82 | 418.74±108.25 | 0.005* |
| Left Superior oblique (SO) | 315.33±99.93 | 267.05±78.92 | 0.083 |
| Left Inferior rectus (IR) | 512.07±98.18 | 398.00±69.46 | <0.001* |
| Left Inferior oblique (IO) | 282.52±82.17 | 215.74±67.51 | 0.005* |

\* denotes significant p-value

In brief, the average EOM volumes (mm^3^) in the left eye of idiopathic HP patients were 466.046 ± 170.261 mm^3^, and in the right eye were 472.022 ± 168.304 mm^3^, while the control group had 373.557 ± 137.288 mm^3^ and 384.296 ± 144.018 mm^3^. The average EOM volumes of both the left and right eyes were significantly greater than those of the control group (p ≤ 0.0001). (Fig. 2)

**Fig. 2.**
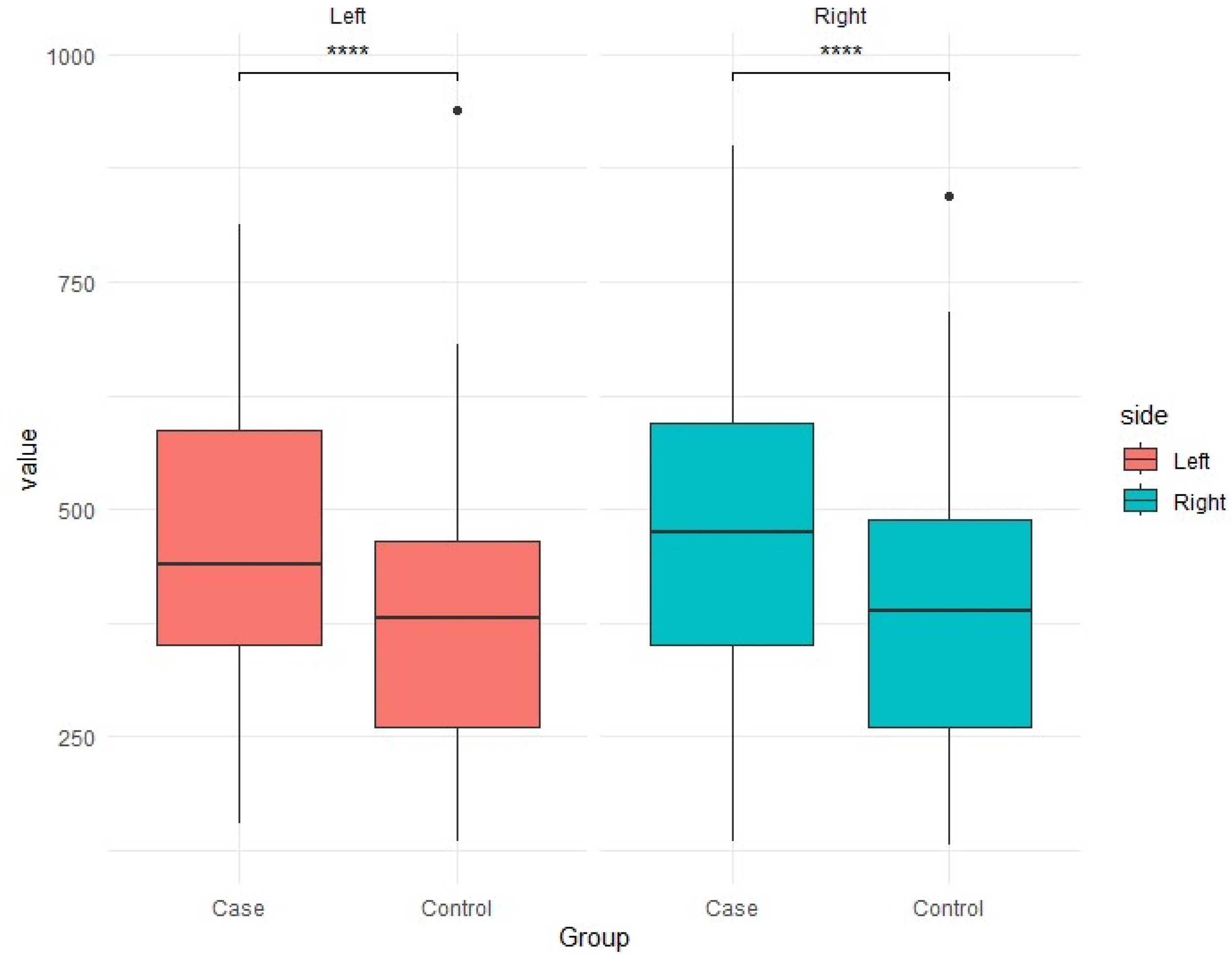
Box plots comparison of the extraocular muscle volume of left and eyes between case (idiopathic HP patients) and healthy controls.

For each EOM, the mean volumes in idiopathic HP patients were 560.65 ± 122.89 mm^3^ in the right medial rectus, 624.03 ± 131.76 mm^3^ in the right lateral rectus, 509.63 ± 154.99 mm^3^ in the right superior rectus, 340.71 ± 70.15 mm^3^ in the right superior oblique, 496.78 ± 134.01 mm^3^ in the right inferior rectus, 300.31 ± 117.56 mm^3^ in the right inferior oblique, 554.06 ± 138.19 mm^3^ in the left medial rectus, 599.05 ± 145.34 mm^3^ in the left lateral rectus, 533.22 ± 146.82 mm^3^ in the left superior rectus, 315.33 ± 99.93 mm^3^ in the left superior oblique, 512.07 ± 98.18 mm^3^ in the left inferior rectus, and 282.52 ± 82.17 mm^3^ in the left inferior oblique muscles.

In comparison to age-matched and sex-matched healthy controls, 9 of 12 EOM muscles showed significantly larger volumes. These included both MR (each p=0.002), both IR (right p=0.08 and left p<0.01), both IO (right p=0.009 and left p=0.005), right LR (p=0.005), right SO (p=0.004), and left SR muscles (p=0.005). However, there was no statistically significant difference in the volume of the right SR, left LR, and left SO muscles (p = 0.096, p = 0.054, and p = 0.083, respectively).

### Relationship between clinical and MRI measures

Correlation with ocular function was assessed only in 9 of the 12 EOM (both MR, both IR, both IO, right LR, right SO, and left SR), which had statistically significant enlargement from prior analysis. Using Pearson’s correlation, there was no statistically significant correlation between all 9 enlarged EOM and ocular motility in idiopathic HP patients, as presented in Table 3.

**Table 3.** Correlation between the significant enlarged EOM volumes (mm3) of idiopathic HP patients and the ocular motility.

| Extraocular muscle | Mean volume in HP (mm<sup>3</sup>), Mean±sd | Percentage of ocular motility limitation ,Mean±sd | Pearson Correlation(P-value) |
| --- | --- | --- | --- |
|  |  |  | EOM volume vs percentage of ocular motility limitation |
| Right IO | 300.31±117.56 | 11.59±24.17 | -0.142(0.527) |
| Right IR | 496.78±134.01 | 11.59±24.94 | -0.337(0.125) |
| Right SO | 340.71±70.15 | 11.59±24.94 | 0.051(0.821) |
| Right MR | 560.65±122.89 | 7.50±21.69 | -0.013(0.955) |
| Right LR | 624.03±131.76 | 26.14±33.37 | 0.249(0.264) |
| Left SR | 533.22±146.82 | 3.18±9.45 | -0.107(0.637) |
| Left IO | 282.52±82.17 | 4.55±13.35 | 0.180(0.423) |
| Left IR | 512.07±98.18 | 3.18±9.45 | -0.221(0.324) |
| Left MR | 554.06±138.19 | 4.55±11.84 | -0.321(0.145) |

A p-value <0.05 indicates statistical significance

## 5. Discussion

Hypertrophic pachymeningitis (HP) is a rare fibrosing inflammatory disorder characterized by thickening of the dura mater, either focal or diffuse. Pathological examination reveals non-specific chronic inflammation, showing lymphoplasmacytic infiltration and dense fibrosis (10). The diagnosis of idiopathic HP is made by excluding various infectious, autoimmune, and neoplastic infiltration diseases. Therefore, when the clinical and imaging findings are consistent, a comprehensive investigation should be conducted to rule out any underlying pathological causes for the condition.

In our study, we observed that 9 out of 12 EOMs (both MR, both IR, both IO, right LR, right SO, and left SR) in idiopathic HP patients were significantly enlarged compared to the healthy control group. There have been no systematic studies specifically comparing extraocular muscles (EOM) in idiopathic HP patients and healthy individuals. However, many studies have been conducted on IgG4-related disease, which is a subset of HP associated with an autoimmune disorder. These studies have reported that the disease can infiltrate the EOM and cause multiple EOM enlargements. IgG4-related disease is a chronic inflammatory disorder characterized by IgG4-positive lymphoplasmacytic infiltrative lesions. Both idiopathic HP and IgG4-RD share similar demographics, histopathology, and natural histories (10-12). Based on its pathophysiology, we hypothesize that idiopathic HP may be a diffuse disease that potentially involves microstructural changes in the EOM, leading to structural volume alterations.

Also, it was noted that there were no studies about predilection muscle involvement in idiopathic HP. In our study, we observed that the left inferior rectus and both medial rectus muscles were significantly larger. As compared with other diseases that cause multiple EOM enlargements, such as IgG4-related disease, the LR muscle is most frequently affected, and in Graves’ ophthalmopathy, the IR, MR, SR, and LR are mostly involved in that order (13-16). However, because our sample size was small, we were unable to conclude the predilection of specific muscles. Further studies may provide more information.

The extraocular movement limitation can occur due to various reasons, including EOM enlargement, like in Graves’ orbitopathy, where it is believed that more EOM enlargement causes a more severe limitation of eye movement (8, 17). Therefore, this study was conducted to clarify whether the restriction of ocular motility is caused by EOM enlargement or not. However, our study found no significant correlation between increased EOM volume and impaired ocular muscle movement in idiopathic HP patients. Previous research suggests the clinical manifestation of idiopathic HP depends on the location of dura infiltration surrounding the adjacent cranial nerves (8, 18). Specifically, deficits in cranial nerves III, IV, and VI are associated with thickened meninges from the cavernous sinus to the superior orbital fissure. Therefore, we imply that the limitation of EOM in patients with idiopathic HP is caused by cranial nerve involvement rather than muscle enlargement.

There are several limitations to this study. Firstly, due to the rarity of idiopathic HP, the single-center study had a small sample size. Secondly, the retrospective nature of the study may affect its reliability. Lastly, although the control patients had an overall physical examination and no history of eye symptoms, they did not undergo a complete ophthalmic examination.

## 6. Conclusion

In idiopathic HP, patients have significantly larger EOM volumes, particularly in the left IR and both MR muscles. This could be due to the diffuse infiltrative histopathology that potentially involves microstructural changes in the EOM. However, no significant correlation was found between the size of these enlarged EOMs and the extraocular movement limitation.

## Data Availability

Data cannot be shared publicly because of confidential patient data. Data are available from the Faculty of Medicine, Siriraj Hospital, Mahidol University Institutional Data Access / Ethics Committee (contact via) for researchers who meet the criteria for access to confidential data.

## Acknowledgements

This research project was supported by the Faculty of Medicine, Siriraj Hospital, Mahidol University.

The authors would like to thank Mr. Karnchana Sae-jang and Mr. Yuti Jirawattanapalin from Siriraj Radiology Department in statistical analysis.

## Author contributions

S.K., S.S. and A.E. collected, assembled, and analyzed data. N.R. and N.C. recruited and examined patients. C.N. performed critical revision of the article for important intellectual content.

## Reference

1. Warittikoon S, Jakchairoongruang K. Distinguishing magnetic resonance imaging features between idiopathic hypertrophic pachymeningitis and secondary hypertrophic pachymeningitis. Asian Biomedicine. 2019;13:113–9. 10.1515/abm-2019-0049

2. Xiao X, Fu D, Feng L. Hypertrophic pachymeningitis in a southern Chinese population: a retrospective study. Frontiers in Neurology. 2020;11:565088. 10.3389/fneur.2020.565088

3. Sakdee W, Termglinchan T, Lertbutsayanukul P. Primary and secondary hypertrophic pachymeningitis in Prasat Neurological Institute: clinical, laboratory and neuroradiologic features. Thai journal of neurology. 2021;37(2):43–57.

4. Charleston Lt, Cooper W. An Update on Idiopathic Hypertrophic Cranial Pachymeningitis for the Headache Practitioner. Curr Pain Headache Rep. 2020;24(10):57. 10.1007/s11916-020-00893-5

5. Yonekawa T, Murai H, Utsuki S, Matsushita T, Masaki K, Isobe N, et al. A nationwide survey of hypertrophic pachymeningitis in Japan. Journal of Neurology, Neurosurgery & Psychiatry. 2013. 10.1136/jnnp-2013-306410

6. Tian S, Nishida Y, Isberg B, Lennerstrand G. MRI measurements of normal extraocular muscles and other orbital structures. Graefe’s archive for clinical and experimental ophthalmology. 2000;238:393–404. 10.1007/s004170050370

7. Bijlsma WR, Mourits MP. Radiologic measurement of extraocular muscle volumes in patients with Graves’ orbitopathy: a review and guideline. Orbit. 2006;25(2):83–91. 10.1080/01676830600675319

8. Lee J-Y, Bae K, Park K-A, Lyu IJ, Oh SY. Correlation between extraocular muscle size measured by computed tomography and the vertical angle of deviation in thyroid eye disease. PLoS One. 2016;11(1):e0148167. 10.1371/journal.pone.0148167

9. El-Kaissi S, Wall JR. Determinants of extraocular muscle volume in patients with Graves’ disease. Journal of Thyroid Research. 2012;2012. 10.1155/2012/368536

10. Wallace ZS, Carruthers MN, Khosroshahi A, Carruthers R, Shinagare S, Stemmer-Rachamimov A, et al. IgG4-related disease and hypertrophic pachymeningitis. Medicine. 2013;92(4):206. 10.1097/MD.0b013e31829cce35

11. Saitakis G, Chwalisz B. The neurology of IGG4-related disease. Journal of the Neurological Sciences. 2021;424:117420. 10.1016/j.jns.2021.117420

12. Kim N, Yang HK, Kim JH, Hwang J-M. IgG4-related ophthalmic disease involving extraocular muscles: case series. BMC ophthalmology. 2018;18(1):1–7. 10.1186/s12886-018-0819-x

13. Mombaerts I, Rose GE, Verity DH. Diagnosis of enlarged extraocular muscles: when and how to biopsy. Current Opinion in Ophthalmology. 2017;28(5):514–21. 10.1097/ICU.0000000000000395

14. Lakerveld M, van der Gijp A. Orbital Muscle Enlargement: What if It’s Not Graves’ Disease? Current Radiology Reports. 2022;10(2):9–19. 10.1007/s40134-022-00392-y

15. Rana K, Juniat V, Patel S, Selva D. Extraocular muscle enlargement. Graefe’s Archive for Clinical and Experimental Ophthalmology. 2022;260(11):3419–35. 10.1007/s00417-022-05727-1

16. Garau LM, Guerrieri D, De Cristofaro F, Bruscolini A, Panzironi G. Extraocular muscle sampled volume in Graves’ orbitopathy using 3-T fast spin-echo MRI with iterative decomposition of water and fat sequences. Acta Radiologica Open. 2018;7(6):2058460118780892. 10.1177/205846011878089

17. Chen YL, Chang TC, Huang KM, Tzeng SS, Kao SC. Relationship of eye movement to computed tomographic findings in patients with Graves’ ophthalmopathy. Acta ophthalmologica. 1994;72(4):472–7. 10.1111/j.1755-3768.1994.tb02800.x

18. Shao W, Tian C, Gao L, Cui B, Shi Q. Rapid bilateral visual loss as the initial clinical manifestation in idiopathic hypertrophic cranial pachymeningitis. Clinical Case Reports. 2022;10(5):e05825. 10.1002/ccr3.5825

